# Long-Term Opioid Use and Emotional, Cognitive, and Sensorimotor Function in Chronic Back Pain

**DOI:** 10.1101/2022.09.14.22279907

**Authors:** Gaelle Rached, Andrew D. Vigotsky, Paulo Branco, Rami Jabakhanji, Thomas J. Schnitzer, A. Vania Apkarian, Marwan N. Baliki

## Abstract

This study examines cognitive, emotional, sensory, and motor function in patients with chronic back pain (CBP) with and without long-term opioid exposure (CBP+O vs. CBP−O) and the influence of race and sex on these outcomes. We recruited 64 CBP+O (mean 7.4 years of opioid use) and 64 matched CBP−O. We collected pain intensity, pain duration, NIH Toolbox outcomes, and measures of opioid use: dosage, duration, blood levels, and withdrawal symptoms. We compared both groups to a normative sample, examined sex and race influences separately, and related CBP+O’s NIH Toolbox outcomes to their opioid use measures.

CBP+O reported greater pain interference and poorer emotional function. Sex moderated opioid exposure’s effects on cognitive performance and social satisfaction, while race moderated motor dexterity and taste outcomes. In CBP+O, 1) opioid dosage was associated with cognitive speed and sensory performance; 2) blood levels of opioids corresponded with poorer attention, motor endurance, and more severe stress and negative affect; 3) opioid use duration was related to improved working memory; while 4) higher withdrawal symptoms were associated with poorer cognitive performance, worsened negative emotions, and decreased grip strength.

The study demonstrates both positive and negative outcomes associated with long-term opioid use in CBP, highlighting the need to consider sex- and race-related factors when assessing function in this population.

## Introduction

The opioid epidemic is a major public health crisis affecting the United States, with devastating health, economic, and societal costs ^1^. Despite controversial evidence of efficacy, opioids are widely used for treating chronic pain and thus contribute to the ongoing epidemic ^2-6^. Chronic pain and opioid use have largely been studied independently compared to healthy controls, although they are intertwined. Chronic pain is associated with impairments in memory, attention, processing speed, and executive functioning ^7, 8^. Similarly, long-term opioid prescription use is associated with declines in global cognitive performance, particularly in memory, language, and attention ^9^. In addition, prescription opioid misuse and chronic pain worsen negative affect and psychological well-being ^10^.

Despite overwhelming evidence that chronic pain and long-term opioid use separately impact multiple psychological domains, insufficient research characterizes both conditions concurrently. Consequently, the characteristics of long-term opioid use in chronic pain patients compared to chronic pain without opioids are poorly understood. Here, we investigated the effect of opioids on patients with chronic back pain (CBP) by comparing CBP on long-term opioids (CBP+O) to matched CBP without opioids (CBP−O) using a standard multidimensional assessment tool, the NIH toolbox, which measures cognitive, emotional, sensory, and motor function ^11^, providing a holistic picture of neurological and behavioral health. As the patients studied were 58% female and 30% black, we also interrogated sex and race influences on NIH toolbox outcomes. Finally, we examined how opioid dosage, opioid blood levels, opioid duration, and withdrawal symptoms related to the NIH toolbox scores in CBP+O.

## Methods

### Recruitment

We recruited 64 CBP+O patients (pain duration > 6 months, mean = 16.5 years) on opioid therapy for at least three months. We selected 64 CBP−O patients (mean pain duration = 16.7 years) from a previously recruited sample (Figure 1). Patients were matched on age, sex, pain intensity, and pain duration using optimal matching ^12^ (Table 1). All participants were fluent in English, over 18 years of age, and could understand instructions and questionnaires. Recruitment was from the Northwestern Medicine (NM) healthcare system and Shirley Ryan Ability Lab. Our exclusion criteria included (1) treatment with a spinal cord stimulator; (2) diagnosis of rheumatoid arthritis, ankylosing spondylitis, acute vertebral fractures, fibromyalgia, low back/spine oncologic history, and other comorbid neurological disorders, including major depression and psychiatric disorder requiring treatment; (3) involvement in litigation regarding their back pain, having a disability claim, or receiving workman’s compensation; (4) significant comorbid diseases such as uncontrolled hypertension, unstable diabetes mellitus, renal insufficiency, congestive heart failure, coronary or peripheral vascular disease, chronic obstructive lung disease, or malignancy; (5) pregnancy; (6) Beck Depression Inventory (BDI) > 28. The protocols were approved by Northwestern University IRB (STU00207384, STU00205398).

**Figure 1:**
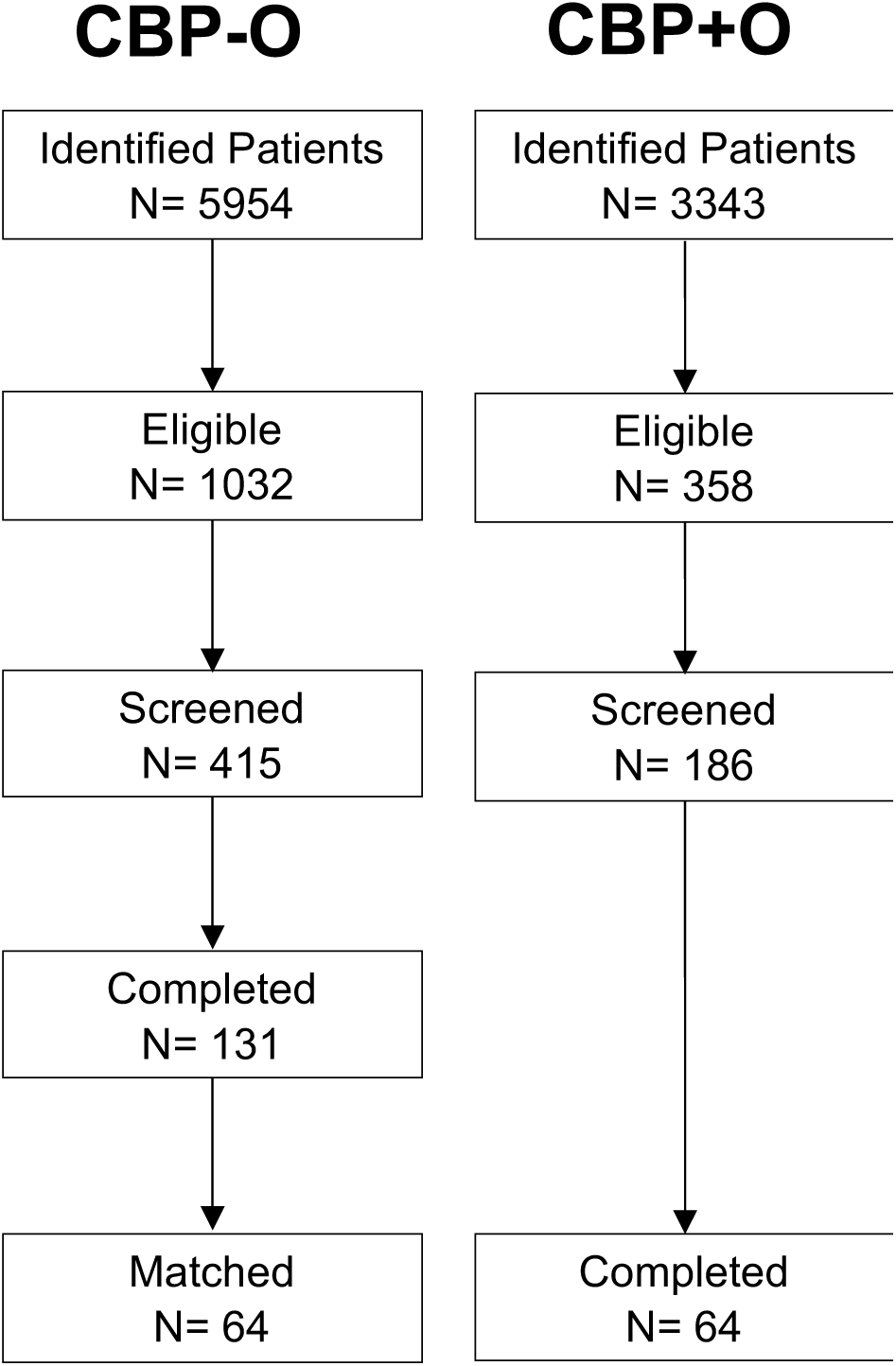
Recruitment flowchart of patients with chronic back pain not consuming opioids (CBP – O) and on long-term opioids (CBP + O).

**Table 1:**
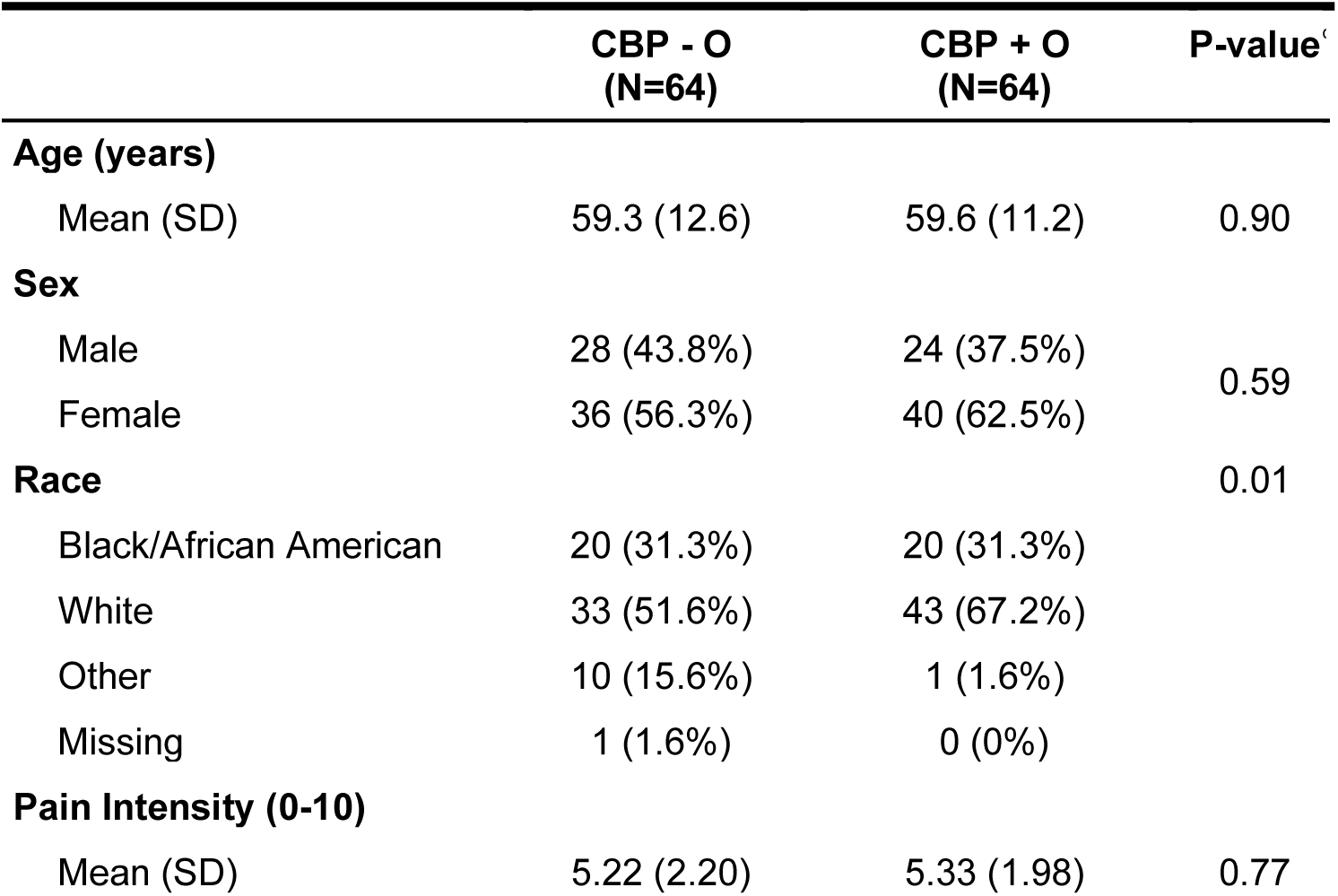
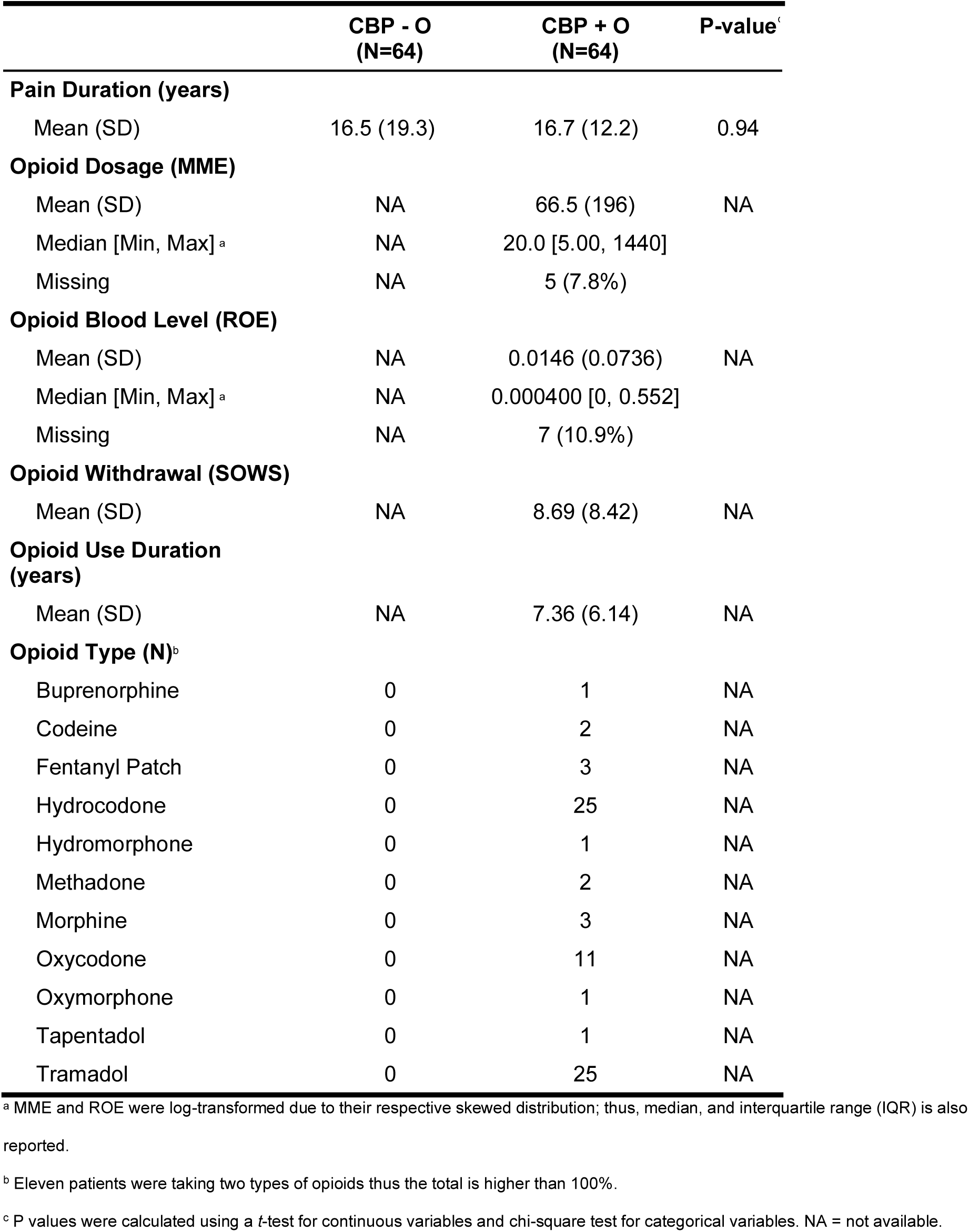
Demographic characteristics of patients with chronic back pain not consuming opioids (CBP – O) and on long-term opioids (CBP + O)

### Measures

We collected demographic information from participants, including age, sex, race, and pain intensity (0–10 numeric rating scale, NRS) and duration. Participants completed the four neuropsychological domains of the NIH toolbox: cognition, emotion, motor, and sensation ^11^. Tests requiring personal interaction between the experimenter and participants were skipped during the height of the COVID-19 pandemic.

For the CBP+O group, we collected opioid duration and dosage, which we converted to morphine milligram equivalent (MME) using CDC’s conversion factors ^13^. We also collected plasma to quantify systemic opioid levels. Plasma opioid concentrations were converted to a relative opiate equivalent (ROE), taking into consideration the affinity of each opioid to the mu-opioid receptor (MOR) ^14^ and respective molar weights ^15-22^. Withdrawal was assessed by the Subjective Opiate Withdrawal Scale (SOWS), a self-report questionnaire to rate common signs and symptoms of opiate withdrawal ^23^.

### Blood sample collection, analysis, and ROE calculations

We also collected blood samples from participants to quantify systemic levels of opioids. The concentrations of eight analytes (oxymorphone, hydromorphone, oxycodone, hydrocodone, fentanyl, buprenorphine, methadone and tramadol) and the presence of three others (morphine, codeine, and heroin) in plasma were determined by liquid chromatography-tandem mass spectrometry after sample preparation by solid-phase extraction, using deuterated analogs (oxymorphone-D3, hydromorphone-D6, oxycodone-D6, hydrocodone-D6, buprenorphine-D4, methadone-D3, and tramadol-C13D3) as internal standards, based on the method reported by Langman et al ^24^. One hundred microliters of the plasma sample were diluted with 300 uL of 1% acetic acid, vortexed and loaded onto a Phenomenex Strata-X-Drug B 96-well plate. After washing with 100 mM sodium acetate buffer and methanol, the analytes were eluted with an ethyl acetate/isopropanol/ammonium hydroxide (7:2:1 v/v/v) mixture and evaporated to dryness under a stream of nitrogen. The samples were reconstituted in 15% methanol in water and analyzed by a Sciex 6500+ Qtrap LC-MS/MS system equipped with a Shimadzu Nexera2 UPLC system. Samples were eluted from a Phenomenex Kinetex 2.6 μm Biphenyl (50 x 2.1 mm) column with a mobile phase consisting of A: water with 0.1 % formic acid and B: methanol with 0.1 % formic acid at a flow rate of 0.6 mL/min with a gradient from 10% B to 85% B in 4.5 min.

Samples were run in duplicate, and the average opioid concentration was used. The opioid concentrations were then converted to a relative opiate equivalent (ROE), taking into consideration the affinity of each opioid to the mu-opioid receptor (MOR) ^14^, and the respective molar weights ^15-22^. We realize that limiting tramadol to its MOR affinity excludes other aspects of the tramadol effect; however, we are specifically interested in the opiate activity on MOR. Samples with peak presence of opioids below the lower limit of quantitation were imputed as the halfway point from zero to the measurable lower quantifiable threshold.

### Statistical Analysis

NIH toolbox differences between CBP+O and CBP−O were analyzed using multiple regression with age, gender, pain intensity, and pain duration included as covariates—the same covariates on which the participants were matched. Because we used one-to-one matching, we calculated cluster-robust variances. For each outcome, each group’s score was also compared to normative data using a one-sample *t*-test, with a mean T-score of 50 and a standard deviation of 10 in the normative sample. To investigate the effects of sex and race, we performed separate multiple regressions: one with a sex-by-group interaction and another with a race-by-group interaction.

To map NIH toolbox scores onto CBP−O’s opioid parameters, we calculated rank-based partial correlations (95% CI) between the NIH toolbox outcomes and measures of opioid use, adjusted for age, sex, pain intensity, and pain duration. To do so, (1) all the measures were converted to ranks, (2) opioid use metrics and NIH toolbox metrics were residualized with respect to the covariates, and (3) we calculated Pearson correlation coefficients using the residuals. Measures of opioid use were quantified in terms of opioid dosage (MME), opioid blood level (ROE), opioid duration (years), and opioid withdrawal (SOWS). MME and ROE were log-transformed due to the skewed distributions.

## Results

We independently evaluated the groups’ NIH toolbox outcomes and contrasted the groups with and without sex and race interactions (figure 2). In the cognition domain, Flanker’s attentional inhibitory control test was most perturbed in both groups compared to normative values, while picture vocabulary was worse in CBP+O. In the emotion domain, CBP+O was worse than CBP−O on all five tests, especially regarding social satisfaction. All six motor tests were abnormal but with divergent differences: balance and walking endurance were similarly diminished in both groups; grip strength was worse in CBP−O; and dexterity was worse in CBP+O. In the sensation domain, taste was similarly worse than normative values across both groups and pain interference was worse in both groups, but more so in CBP+O.

**Figure 2:**
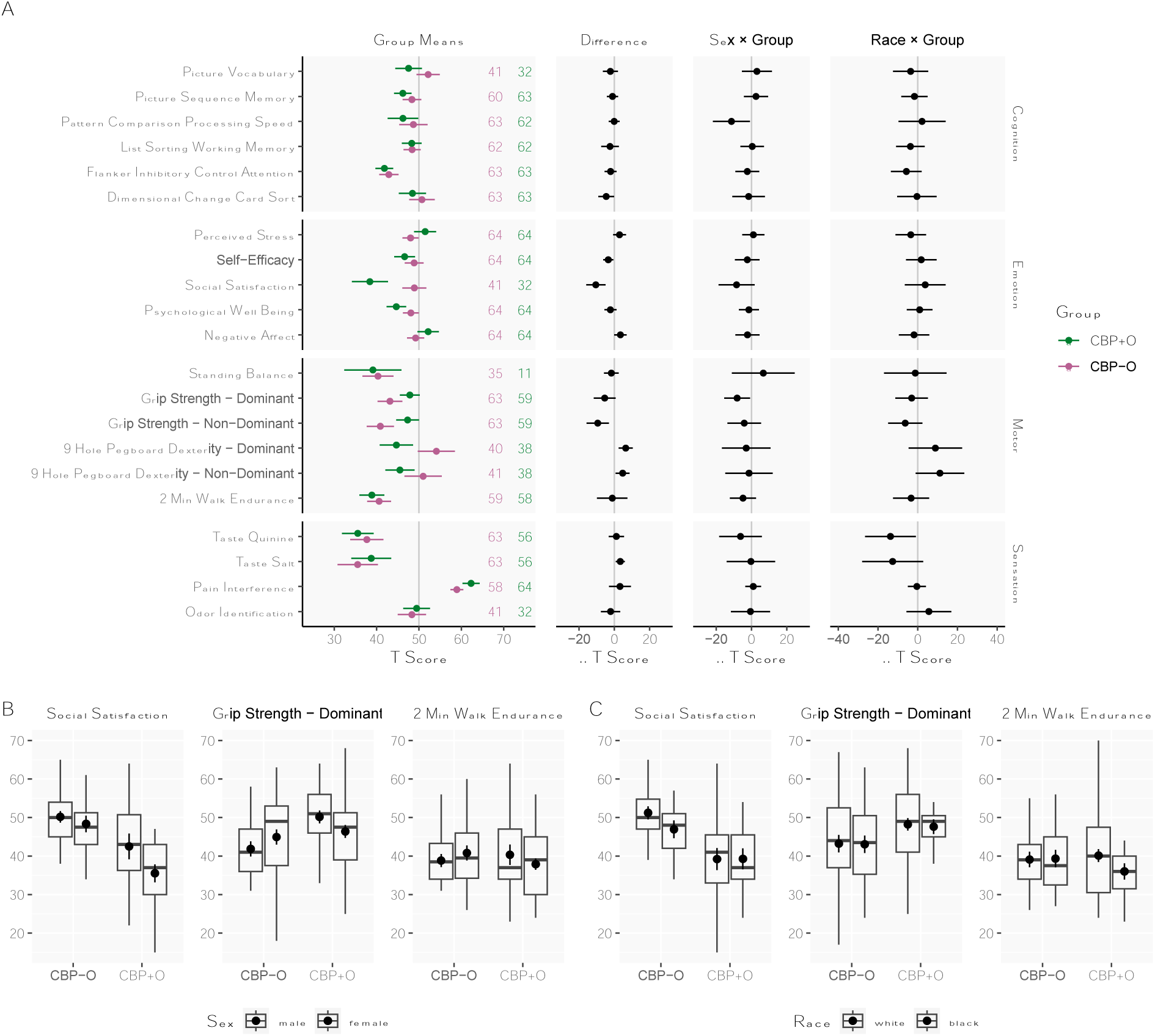
The Emotional, Cognitive, and Sensorimotor Scores in Patients with Chronic Back Pain with (CBP+O) or without Long-Term Opioid Use (CBP-O) and Their Relationships with Sex and Race. (**A**) Estimated marginal means (95% CI) of the NIH toolbox components for patients with chronic back pain with (green, *n*=64) and without (pink, *n*=64) long-term opioids. Point estimates and their CIs were obtained using a linear regression adjusted for age, sex, pain intensity, and pain duration with cluster-robust variances. The grey line indicates the mean in a normative sample (normative T-score = 50 ± 10, n =1000). Each group can thus be directly compared to the normative sample using their estimate and 95% CI. The number of participants per group per measure is shown. The second column shows group differences (CBP+O minus CBP-O, 95% CI) for each subscale. Data points with CIs not crossing the line on 0 indicate *P* < 0.05. The third column shows opioid group by sex interactions (95% CI). Positive values indicate that the CBP+O minus CBP**−**O is greater in females, and negative values indicate the opposite (CBP+O minus CBP**−**O is greater in males). The fourth column shows the opioid group by race interaction (95% CI); positive values indicate that CBP+O minus CBP**−**O is greater in blacks, and negative values indicate the opposite (CBP+O minus CBP**−**O is greater in whites). We did not include the “Other” category for race in the analysis. (**B**) Group differences in Grip-Strength Dominant, Social Satisfaction, 2min Walk Endurance stratified by sex. (**C**) Group differences in Grip-Strength Dominant, Social Satisfaction, 2min Walk Endurance stratified by race. Box plots show the upper and lower quartiles and medians, and point estimates show the mean.

Sex moderated the effects of opioid exposure on social satisfaction, grip strength, and walking endurance. Race moderated the group effect on grip strength, dexterity, stress, working memory, and flanker inhibitory control.

Adjusted partial correlations within the CBP+O group examined the influence of opioid use measures on NIH toolbox outcomes (Figure 3): 1) Higher opioid use dosages were related to better processing speed, salt and odor identification, and worse dexterity. 2) Greater opioid blood levels were related to higher stress, decreased well-being, increased negative affect, and diminished walking ability. 3) Longer durations of opioid use were related to better memory performance. 4) More severe opioid withdrawal signs (SOWS) were related to worse vocabulary and memory, worse outcomes for all five emotion measures, lower grip strength and quinine taste, and increased pain interference.

**Figure 3:**
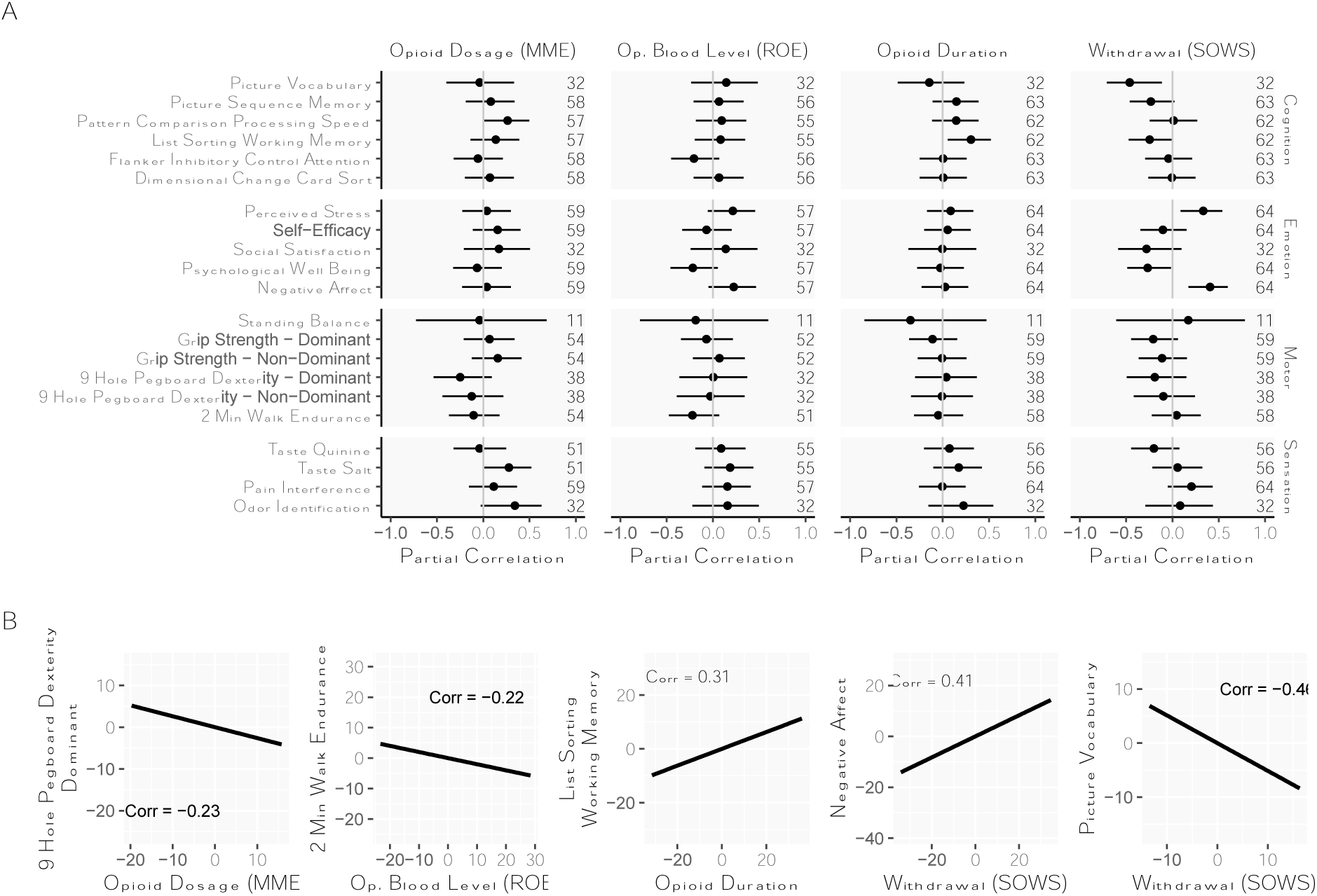
The Relationship between Emotional, Cognitive, and Sensorimotor Scores and Opioid Dosage, Opioid Blood Level, Opioid Use Duration, and Withdrawal Signs in Patients with Chronic Back Pain with Long-Term Opioid Use (CBP+O). (**A**) Partial rank correlations (95% CI) between the NIH Toolbox items and measures of opioid use adjusted for age, sex, pain intensity, and pain duration. Measures of opioid use are quantified in terms of opioid dosage (log(MME)), opioid blood level (log(ROE)), opioid duration (years) and opioid withdrawal (SOWS). The numbers of participants per NIH Toolbox test and per opioid use measure are represented on the plot. (**B**) Scatter plots showing the partial rank correlations between some NIH Toolbox items and measures of opioid use. Nine-Hole Pegboard Dexterity-Dominant was correlated with opioid dosage (log(MME)), 2-Minute Walk Endurance with opioid blood level (log(ROE)), List Sorting Working Memory with opioid duration, and Negative Affect and Picture Vocabulary with Withdrawal Severity (SOWS). Solid lines in the scatter plots show fitted lines and shaded areas are 95% confidence intervals. Since we used partial rank correlations, the axes in (B) are based on residualized ranks rather than original units.

## Discussion

Our study examined the association of chronic pain and long-term opioid use on cognitive, sensory, emotional, and motor functioning. Both CBP+O and CBP−O experienced more pain interference than the normative sample. Although our study groups were matched on back pain intensity and duration, CBP+O exhibited more pain interference, which we surmise may be related to opioid-induced hyperalgesia ^25, 26^; these effects were not moderated by race and sex. Within CBP+O, pain interference was most strongly related to withdrawal signs, weakly related to opioid dosage (consistent with earlier data ^27^) and opioid blood levels, and appeared to be unrelated to the duration of opioid use (contrary to an earlier report ^28^).

The emotional domain of the NIH toolbox was the dimension most perturbed by opioid exposure. All five emotion-related tests exhibited poorer outcomes in CBP+O, especially regarding social satisfaction, which was moderated by sex. Like pain interference, all five measures were associated with withdrawal and less consistently with blood levels of opioids. Given that this is a cross-sectional observational study, causal relationships remain unclear. However, given that pain interference and emotion domain were all related to state measures (blood opioids, withdrawal), it is suggested that the worsening of these outcomes is a consequence of opioid exposure.

In the motor domain, some measures were improved, and others were exacerbated in CBP+O. Moreover, these outcomes were differentially moderated by race. In the sensory taste outcomes, both CBP−O and CBP+O showed similar abnormalities moderated by race. In the cognitive domain, CBP+O and CBP−O performed poorly on attentional tasks, consistent with previous studies demonstrating that chronic pain is associated with impaired general cognitive functions ^8, 29, 30^. These results have broad implications for the daily functioning and rehabilitation of individuals with chronic pain. Previous results regarding long-term opioid use and cognition remain contradictory and inconclusive ^31-36^.

Disparities in cognitive performance among individuals with chronic pain relative to race highlight the need for further investigation and consideration of race-related factors when assessing and treating cognitive impairments in this population. Interestingly, sex effects were less prominent than race and included specific outcomes in cognition, emotion, and motor domains. Both factors remain to be studied more thoroughly in the future.

This study has three noteworthy limitations. First, our cross-sectional design makes the causality unclear. However, this is partially overcome by our matching and relating outcomes to opioid measures. Second, we excluded patients who have a clinical diagnosis of depression and have a BDI > 28, decreasing the generalizability of our results while allowing us to characterize behavior and functioning in patients with minimal psychiatric confounds. Third, our matched sample comes from a previously recruited sample. Since the data were collected by different people at different time points, potentially affecting our measured values.

## Conclusion

Our study provides important insights into the impact of chronic pain and long-term opioid use on various domains of functioning. Chronic pain was associated with cognitive, sensory, emotional, and motor impairments, with attention deficits being a prominent feature. Our results suggest that long-term opioid use may not significantly contribute to cognitive impairments beyond chronic pain itself. However, long-term opioid use is associated with worse emotional functioning. Additionally, our study highlights potential race and sex disparities in cognitive performance and social satisfaction, respectively, among individuals with chronic pain. Overall, our study furthers our understanding of the complex interactions between chronic pain and long-term opioid use in different aspects of patients’ lives, which can inform more effective treatment strategies for this patient population.

## Data Availability

All data produced are available online at openpain.org

https://www.openpain.org

## Acknowledgments

This work made use of the Mary Beth Donnelley Clinical Pharmacology Core at Northwestern University, which has received support from the NIH (1S10OD012016-01 / 1S10RR019071- 01A1), Soft and Hybrid Nanotechnology Experimental (SHyNE) Resource (NSF ECCS- 1542205), the State of Illinois, and the International Institute for Nanotechnology (IIN).

Meghan Ford, data collection

Elizabeth Yan, data collection

Maryam Abdallah, data collection

Byron K Yip, data collection

Santiago Espinosa, data collection

Rahia Parveen Shuaib, data collection

